# Evaluation of a clinical decision support tool for matching cancer patients to clinical trials using simulation-based research

**DOI:** 10.1101/2021.05.30.21257718

**Authors:** Clarissa Gardner, Jack Halligan, Gianluca Fontana, Roberto Fernandez Crespo, Matthew Prime, Chaohui Guo, Okan Ekinci, Saira Ghafur, Ara Darzi

**Author notes:** **Corresponding author** Name: Clarissa Gardner, Email Address, Address: Institute of Global Health Innovation, Imperial College London, London, UK.

## Abstract

Simulation-based research (SBR) methods have been proposed as an alternative methodology for evaluating digital health solutions; however, applicability remains to be established. This study used SBR to evaluate a clinical decision support (CDS) tool used for matching cancer patients to clinical trials. 25 clinicians and research staff were recruited to match 10 synthetic patient cases to clinical trials using both the CDS tool and publicly available online trial databases. Participants were significantly more likely to report having sufficient time (*p* = 0.020) and to require less mental effort (*p* = 0.001) to complete trial matching with the CDS tool. Participants required less time for trial matching using the CDS tool, but the difference was not significant (*p* = 0.093). Most participants reported that they had sufficient guidance to participate in the simulations (96%). This study demonstrates the use of SBR methods is a feasible approach to evaluating digital health solutions.

## Introduction

The term ‘digital health solution’ describes the use of information and communication technology in healthcare contexts^1,2^. Other commonly used terms include digital health interventions^1^ and digital health technologies^3^. Digital health solutions can include electronic health records, wearable devices and sensors, health analytics, medical imaging, mobile health (mHealth) as well as digital devices and robotics used in healthcare settings^4^. While digital health solutions have been shown to address healthcare challenges in various disease areas and healthcare contexts, there are various implementation challenges. Examples include policy barriers^4,5^, financial barriers^6^, cultural resistance amongst healthcare professionals^7–9^, and the ability to provide timely and robust evidence for the efficacy of digital health solutions^7^.

To justify their implementation into clinical settings and ensure public trust and patient safety, digital health solutions must be evaluated for usability, acceptability, clinical effectiveness and cost-effectiveness^9–11^. Randomized controlled trials (RCTs) have traditionally been recognized as the gold standard to demonstrate the clinical effectiveness of novel interventions and therapeutics^12^. However, few digital health solutions have been evaluated using RCTs, possibly due to the fast-paced nature of innovation and the arduous process of setting up an RCT in a clinical setting^8,12^. The relatively high costs and resources associated with conducting an RCT reduces the return on investment and is therefore not a sustainable option for small and medium-sized enterprises (SMEs) or products in the early stage of development^8^. Furthermore, recruiting participants to RCTs and pilot studies to evaluate digital health solutions faces challenges such as data privacy and security concerns, the commitment required to participate in longitudinal studies for self-management interventions, and a lack of endorsement for trial participation from treating clinicians^8,13–15^.

The regulation and publication of guidance to evaluate digital health solutions is outpaced by the rapidly evolving digital health landscape and the increasingly diverse types of solutions entering the market^16,17^. However, there is a lack of clarity regarding what evidence generation methodologies are appropriate to evaluate solutions in the early stages of product development. In particular, what is needed are evidence generation methodologies that can be applied to various risk categories of digital health solutions that can generate high-quality insights, in a short timeframe, and at relatively low cost^3,8^. A perspective piece published in npj Digital Medicine on the challenges of evaluating digital health solutions proposed alternative methodologies that facilitate safe and responsible growth across the sector^8^, such as the use of simulation-based research (SBR) methods.

### Existing applications of simulation-based research (SBR) methods in healthcare contexts

Simulation is described as the replication of real-life events in an interactive manner, and has been applied in research and the trial of new healthcare interventions^18^. Simulation has also been widely applied to healthcare in the context of education and clinical rehearsal, and allows healthcare providers the opportunity to learn new protocols and procedures in a safe and controlled environment^19–24^.

SBR in the context of healthcare involves replicating clinical scenarios, with the aim of eliciting reactions and behaviors from healthcare professionals that are as close as possible to how they would react in a real situation^25^. SBR may take place in situ (i.e., in a clinical setting), where they can be announced or unannounced (i.e., participating staff are aware that a simulated event will take place but may or may not be informed as to when), or in dedicated simulation spaces away from the clinical setting^26–28^. The design of such environments should take into account the layout, noise isolation and the relatedness to the real-life context^29^.

### Knowledge gap: the feasibility of using SBR methods to evaluate digital health solutions

The use of SBR methods could support faster evidence generation of the usability, acceptability and effectiveness of digital health solutions without involving real patients and/or real patient data. Such simulations could take place outside of a clinical setting, providing opportunities to obtain behavioral and cognitive measures that would otherwise be impractical to gather in routine practice^8^. SBR methods have been used to evaluate healthcare interventions^30,31^, however, few studies have reported the use of SBR methods to evaluate digital health solutions and the results are seldom reported ^8,32–34^.

### Filling the knowledge gap: using SBR methods to evaluate a clinical decision support tool

To assess the feasibility of using SBR methods to evaluate digital health solutions we sought to apply these methods to evaluate a tool which primarily supports existing clinical services yet has no direct impact on patient outcomes. According to guidelines published by the National Institute for Health and Care Excellence (NICE), such solutions would be functionally classified as “Tier 1” and require the minimum standards of evidence for effectiveness^3^. The intended outcome would be to demonstrate that SBR methods could be used in the first instance to generate the minimum level of evidence required to support the adoption or larger-scale evaluation of any digital health solution. The digital health solution chosen for this study was NAVIFY Clinical Trial Match (NAVIFY CTM). NAVIFY CTM is an application within NAVIFY Tumor Board (Roche Information Solutions Inc., Santa Clara, CA)) (NAVIFY TB), a cloud-based workflow solution that is used to prepare for and conduct cancer multidisciplinary team meetings.

NAVIFY CTM is a clinical decision support tool that has been developed to streamline the process of searching for and identifying clinical trials by using patient-specific data such as cancer stage, genomic alterations, and the treating institution’s postcode to perform an automated search for suitable clinical trials across 21 international trial databases. Clinical trials present an opportunity for cancer patients who meet specific trial eligibility criteria to benefit from the therapeutic effects of novel treatments and support the development of treatments that could be implemented in the future^35^. However, while the number of open cancer clinical trials and enrolled participants are increasing^36^, most trials available to cancer patients are under-recruiting^37^. The barriers to recruitment from the patient’s and clinician’s perspective are well-recognized and include challenges identifying eligible patients and approaching them to discuss participation^38^. Evidence suggests that decision-making around clinical care and patient outcomes can be cognitively challenging for clinicians^39,40^ which may contribute to an insufficient number of eligible patients being identified for clinical trials. Therefore, the evaluation of NAVIFY CTM called for methods to generate evidence for its effectiveness in streamlining the process of identifying suitable trials as well as reducing the mental effort for clinicians. It also presented as an opportunity to conduct usability testing with practicing clinicians to provide recommendations for product development and future implementation.

### Aims and objectives of this study

The aims this study are:

- To demonstrate that SBR methods are a viable approach to evaluate clinical decision support tools.
- To use SBR to evaluate NAVIFY Clinical Trial Match (Roche Information Solutions Inc., Santa Clara, CA)) herein referred to as NAVIFY CTM, a clinical decision support (CDS) application designed to match patients to clinical trials.

The objective of this study is to evaluate the efficiency, quality, and cognitive burden of using NAVIFY CTM to match patients to cancer clinical trials, with publicly available online trial databases used as the comparator, for example, ClinicalTrials.gov.

## Methods

### Study design and participants

We performed an experimental within-subject, non-blinded study to evaluate NAVIFY CTM using simulation based-research (SBR) methods. Participants were approached as previous participants of digital health studies conducted at the university or via the research team’s professional networks. Snowball sampling was used in this study to reach additional participants. This method is a commonly used recruitment technique in qualitative research when trying to engage more individuals in a specific population^41^. 25 clinicians were recruited to participate in the simulation sessions, all of whom had experience of matching patients to cancer clinical trials in the UK. Participants included consultant oncologists (i.e., board certified attendings), specialist registrars in oncology (i.e., senior residents), research nurses and clinical trial practitioners.

The research team identified 10 actively recruiting Investigational Medicinal Product (IMP) trials that represented a mix of trials for small cell lung cancer (SCLC) and non-small cell lung cancer (NSCLC). A total of 11 synthetic patient cases were created to match each of these trials (two of the cases were eligible for the same trial). The cases were developed by two consultant thoracic oncologists, one consultant histopathologist, and one consultant interventional radiologist, and included clinical details (e.g., past medical history, social history), pathology reports, and radiology reports with accompanying images. The patient cases were added to NAVIFY TB to facilitate trial matching using the NAVIFY CTM application. Online trial databases that are known to be in use in clinical practice were used as the study comparator. These databases are publicly accessible and are designed to enable members of the public, clinicians and research staff to identify clinical trials. To facilitate trial matching using online trial databases, clinical details and reports were provided on a laptop computer in PDF format with images provided in JPG format.

Participation in this study was pseudonymous and ethics approval for this study was sought from and approved by the Research Governance & Integrity Team at the university (ICREC reference: 19IC5457). Informed consent was obtained from participants at the beginning of each simulation session. The study was funded by F. Hoffman-La Roche Ltd, and participants were reimbursed for their time and travel expenses.

### Study procedures

Participants were instructed to assume the role of a clinician or research staff member at St Mary’s Hospital in London who is preparing to see 10 patients at an outpatient clinic. Participants were advised to find at least one suitable clinical trial for each patient with the intention of discussing trial enrolment and to treat the session as they would a real-life clinical scenario.

Table 1 shows the order of procedures during the simulation sessions. Each participant completed two separate trial matching exercises – one exercise matching trials for five of the 10 patients using NAVIFY CTM, and one exercise matching trials for the remaining five patients using publicly available online trial databases. The order of the trial matching exercises was randomly assigned so that half of the participants started with NAVIFY CTM and half started with online trial databases. The synthetic patient cases were also randomly assigned. All trial matches were recorded by participants in Microsoft Excel. Participants were also required to complete an online Stroop Color and Word Test (SCWT) before and after each trial matching exercise. The SCWT measures participants’ reaction times and the accuracy of their response to text color and wording pairs (a ‘Stroop task’) each of which are either congruent between text and color (e.g., “GREEN”) or incongruent (e.g., “RED”). The SCWT has been used pre- and post-task to measure the mental agility of study participants who have completed mentally demanding activities^42^. The SCWT has been used to measure conflict-controlling selective attention/response inhibition and working memory^43,44^.

**Table 1.** The order of procedures during the simulation sessions.

| Activity | Duration (mins) |
| --- | --- |
| Informed consent and introductions | 5 |
| Review of simulation session guide | 10 |
| Practice Stroop Color and Word Test (SCWT) | <1 |
| SCWT 1* | <5 |
| Trial matching exercise 1** | 60 |
| SCWT 2* | <5 |
| BREAK | 10 |
| SCWT 3* | <5 |
| Trial matching exercise 2** | 60 |
| SCWT 4* | <5 |
| BREAK | 10 |
| Post-simulation survey | 10 |
| **Total** | **~180** |

\* Participants completed the Stroop Color and Word Test (SCWT) before and after each trial matching exercise. This was used to infer participants' mental agility before and after each trial matching exercise and estimate the cognitive burden associated with matching the synthetic patient cases using NAVIFY CTM or online trial databases.
\*\* Participants were randomly assigned to complete the trial matching exercise with NAVIFY CTM or online trial databases first.

Before the NAVIFY CTM trial matching exercise, participants were given a brief demonstration of how to navigate NAVIFY TB and how to access NAVIFY CTM. Participants were also provided with a printed copy of the NAVIFY CTM User Guide and shown a 5-minute video on how NAVIFY CTM is used. For the comparator, participants were provided with a list of links to online trial databases that are used in clinical practice. Participants were free to use one or multiple of the below databases to find a suitable trial. The online trial databases used in the study included ClinicalTrials.gov^45^, EU Clinical Trials Register^46^, NIHR CRN Portfolio Search website^47^, NIHR ‘Be Part of Research’ website^48^, International Standard Randomized Controlled Trial Number (ISRCTN) registry^49^, and North West London Clinical Research Network (NWL CRN) Lung Cancer Portfolio^50^.

A simulation session guide was developed by the research team to describe the simulated clinical scenario. Participants were instructed to read this document at the beginning of each session. A member of the research team was present throughout each session to resolve technical issues and to ensure data collection procedures were adhered to.

To reflect the real-life time pressure of a clinician tasked with this responsibility, participants were given 60 minutes for each of the two trial matching exercises (i.e., an average of 12 minutes per patient). All trial matches were recorded by participants in Microsoft Excel. The simulation session concluded with an online Qualtrics survey. Questions in the survey were informed by a review of the literature related to clinical trial matching in the UK and by interviews conducted with oncologists and research staff. When all study procedures were accounted for, each simulation session lasted approximately three hours.

### Outcomes

#### Trial matching efficiency

The start time of each trial matching exercise was recorded by a member of the research team. Participants were required to record the time they finished each case in Microsoft Excel. Screen recording software Snagit was used to record the computer screen during the simulation sessions. A web browser plug-in called Webtime Tracker was used to record the amount of time participants spent on each website during the sessions.

#### Quality of trial matches

The trial matches identified by participants were independently scored by a consultant thoracic oncologist. Trials were scored from 0 to 5 based on the following criteria: 0 – No eligibility criteria matched; 1 – Age, tumor type and location matched; 2 - Age, tumor type, location and TNM staging matched; 3 - Age, tumor type, location, TNM staging, performance status, RECIST criteria matched; 4 - Age, tumor type, location, TNM staging, performance status, RECIST criteria, previous lines of treatment matched; 5 - Age, tumor type, location, TNM staging, performance status, RECIST criteria, previous lines of treatment, biomarkers and all other eligibility criteria met.

#### Cognitive burden

The SCWT were programmed using Psytoolkit.org, with each SCWT containing 40 individual Stroop tasks. The maximum response time of each Stroop task was limited to 1.5 seconds. If no response was detected within the response time, this task was considered an error. The post-simulation survey included a subjective measure of mental effort via the Paas scale^51^ which is a popular measure of cognitive load and has been used to validate other measures of cognitive load^52^.

#### Using SBR to evaluate digital health solutions

Participants completed a survey after the simulation session and were invited to provide feedback on the usability and potential applications of NAVIFY CTM in clinical practice, as well as their experience of participating in the simulation sessions and how realistic the synthetic patient cases were.

### Statistical analysis

The scores for the trial matches and the data from the SCWTs, Webtime Tracker, and survey were compiled using Microsoft Excel. Free-text responses to the post-simulation survey underwent thematic analysis^53^ using an inductive approach whereby the onus to derive themes was placed on the researchers. Data was imported into and analyzed on R version 4.0^54^.

All outcome variables used in this analysis were non-normally distributed, as confirmed by Shapiro-Wilk test results. For assessment of trial matching efficiency, outliers were first identified using Ronser’s test (EnvStats package, version 2.4)^55^. This method was selected as it allows simultaneous identification of multiple outliers within sufficiently large datasets.

Trial matches over 30 minutes both belonging to matches found using the ClinicalTrials.gov database) were identified as outliers and subsequently removed from the analysis.

After completing the trial-matching exercise, participants were asked to rank their sense of having sufficient time to complete the task, and the relevance of the suggestions provided by each database. These questions were structured in a Likert scale with the following possible answers: Completely agree, somewhat agree, neither agree nor disagree, somewhat disagree, completely disagree. Answers to these questions were converted into a one (Completely disagree) to five (Completely agree) scale for analysis. Quality of the trials found by the participants of the study was scored independently by a thoracic oncologist, rating all matches in a one (worst possible match) to five (best possible match) scale. Cognitive burden was measured in a Paas scale, and converted to a one (very, very low mental effort) to nine (very, very high mental effort) scale. All pairwise comparisons were performed using a Wilcoxon Signed-Rank test, and *p* values were adjusted using the false discovery rate (FDR) method due to the small number of comparisons being performed at each step.

## Results

### Using SBR to evaluate digital health solutions

#### Feedback on simulation sessions and synthetic patient cases

The majority of participants (n = 24, 96%) stated that they were provided with sufficient guidance to participate in the simulations and complete the trial matching exercises. Most participants (n = 18, 72%) also reported that they were provided enough clinical information in the synthetic patient cases. A small number of participants said that it would have been beneficial to have more histology information (n = 2, 8%), more information on previous treatments (n = 2, 8%), and the inclusion of clinical information via a referral letter (n = 1, 4%).

#### Feedback on the use of simulation-based research (SBR) methods

Table 2 shows the themes derived from the free-text responses in the post-simulation survey on the benefits and challenges of using SBR to evaluate digital health solutions. Benefits identified by participants included the opportunity for clinicians to preview solutions before they are implemented in clinical practice (n = 9, 36%) and to provide feedback on early iterations of digital health solutions (n = 7, 28%). Challenges reported by participants included a lack of familiarity with the novel solution (n = 3, 12%) and the time constraints to complete the relevant exercises (n = 2, 8%). A small number of participants also stated that simulation sessions could be challenging to run if the patient cases lack sufficient data (n = 2, 8%) or if the simulations took too much time away from clinical duties (n = 2, 8%).

**Table 2.** Analysis of free-text responses to the post simulation survey regarding the use of simulation-based research methods to evaluate digital health solutions.

|  | n, (%) |
| --- | --- |
| ***Simulation Benefits*** |  |
| Preview of app in real-time/experience before launch | 9, 36% |
| Provide amendments/feedback before app launch | 7, 28% |
| Broad staff perspective during testing | 3, 12% |
| Training/familiarity before use in clinical practice | 3, 12% |
| ***Simulation Challenges*** |  |
| Additional time/training required to learn how to use search tools | 5, 20% |
| Lack of time for a challenging exercise | 4, 16% |
| More clinical information available in a real scenario | 2, 8% |
| Participant selection bias / personal limitations | 2, 8% |
| Small sample size of participants/cases | 2, 8% |
| Taking time from clinical duties to participate | 2, 8% |
| WiFi connectivity during simulation | 2, 8% |
| Feeling rushed to complete trial matching exercises | 2, 8% |
| Small range of cases/tumor types | 2, 8% |
| Length of simulation session should be shorter | 1, 4% |
| Unable to use real hospital data | 1, 4% |
| Participant lack of familiarity with lung cancer | 1, 4% |
| Inability to get feedback from clinical team on eligibility | 1, 4% |
| Difficult to simulate institutional logistics | 1, 4% |

### Evaluating NAVIFY Clinical Trial Match

#### Participant characteristics

A total of 25 clinicians and research staff with experience screening patients for clinical trials in oncology participated in the study. All participants had experience screening patients in public teaching hospitals operated by the National Health Service (NHS) in the UK. Roles of participants included clinical trial practitioners (*n* = 8, 32%), research nurses (*n* = 8, 32%), specialist trainees in oncology (*n* = 7, 28%) and consultant oncologists (*n* = 2, 8%). Approximately half of participants (*n* = 12, 48%) had greater than five years’ experience screening patients for clinical trials in oncology and half had less than five years’ experience (*n* = 13, 52%).

#### Overview of trial matches

Each of the 25 study participants were asked to match 10 synthetic patient cases to lung cancer clinical trials – five cases using NAVIFY CTM and five using online trial databases – and participants could match a case to more than one trial. A total of 342 trial matches were made across the 10 synthetic patient cases. Of these, 172 trial matches were made using NAVIFY CTM and 170 using online trial databases. Participants failed to match a trial for 34 cases (14%), with 14 of these 34 failures occurring while using NAVIFY CTM and 20 while using online trial databases. Failing to match a case to a trial was not linked to a specific patient case, with six case match failures being the highest for any patient case. When using online trial databases, ClinicalTrials.gov was used for the majority of trial matches (71%) followed by the North West London CRN Lung Cancer Portfolio (12%).

#### Trial matching efficiency

Participants matched trials faster on average when using NAVIFY CTM (10 minutes) compared to the average across all online trial databases (11.7 minutes), although the difference was marginally non-significant (*p* = 0.063) and results stayed similar when outliers (*n* = 2) were removed from the dataset (*p* = 0.092) (Figure 1). The time taken to match the synthetic patient cases to clinical trials using NAVIFY CTM and each of the online databases used by participants in this study is summarized in Table 3.

**Table 3.**
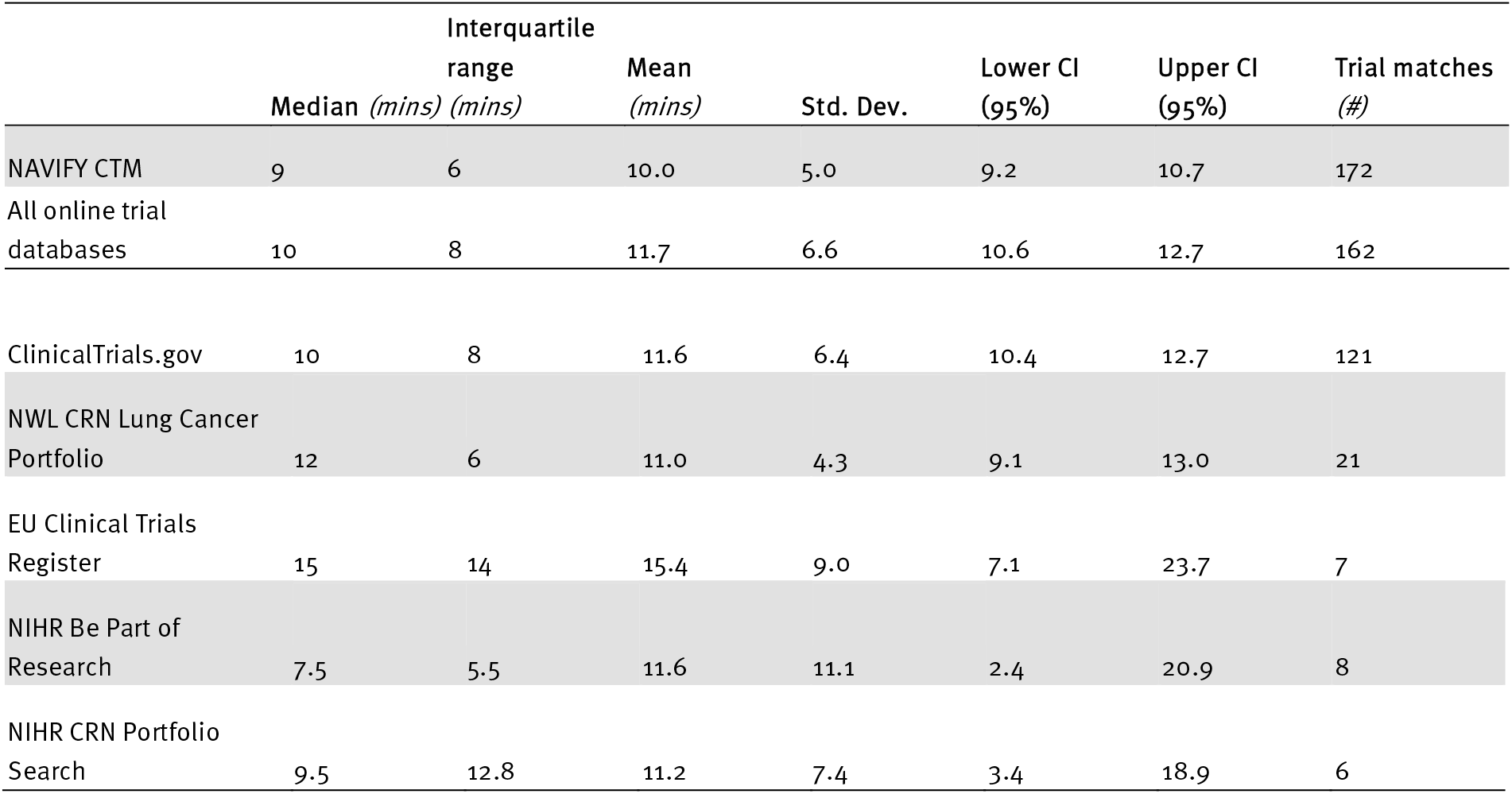
Summary of time to match patient cases to clinical trials (NAVIFY CTM vs. online trial databases).

**Figure 1.**
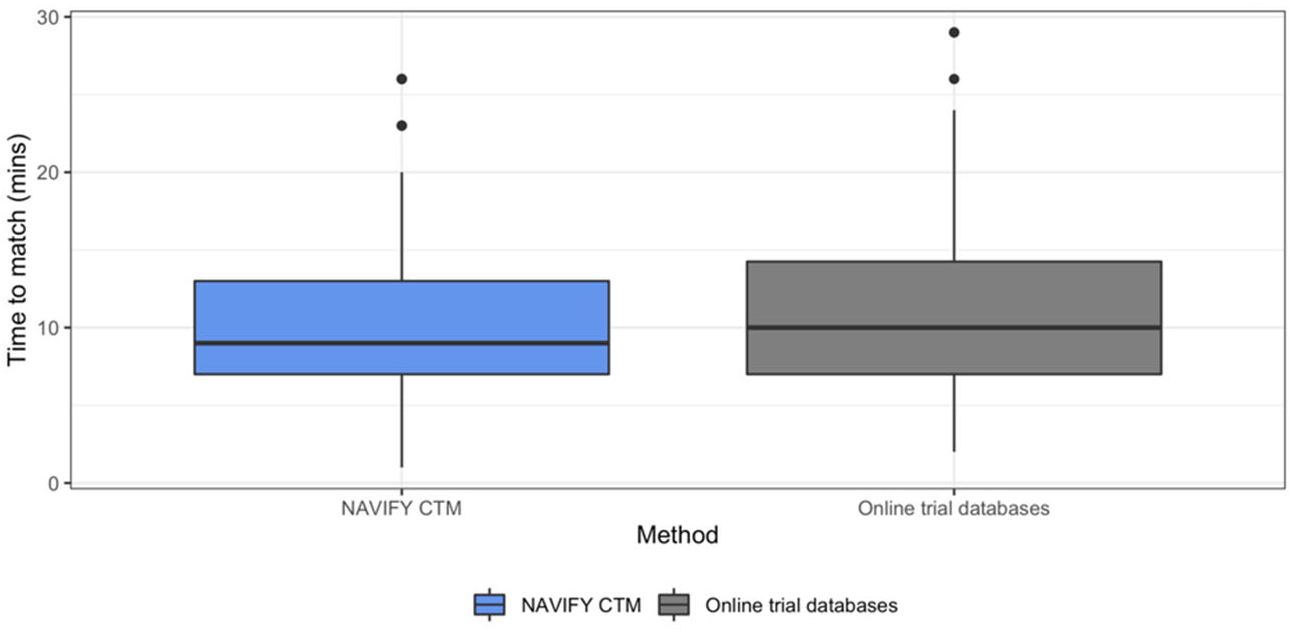
Time to match patient cases to clinical trials, NAVIFY CTM vs. all online trial databases combined (Outliers illustrated as black dots and removed in the statistical tests). Participants matched trials faster using NAVIFY CTM, although the difference was not statistically significant (p = 0.092).

In the post-simulation survey, we identified significant differences when comparing the perception of having sufficient time to match patients to clinical trials (Figure 2). Participants reported a significantly higher degree of agreement with having sufficient time to complete the trial matching exercise using NAVIFY CTM (*p* = 0.020). There was a larger proportion of individuals indicating they ‘completely agree’ that they had enough time to complete the task using NAVIFY CTM (n = 14, 56%) compared to online trial databases (n = 6, 24%).

**Figure 2.**
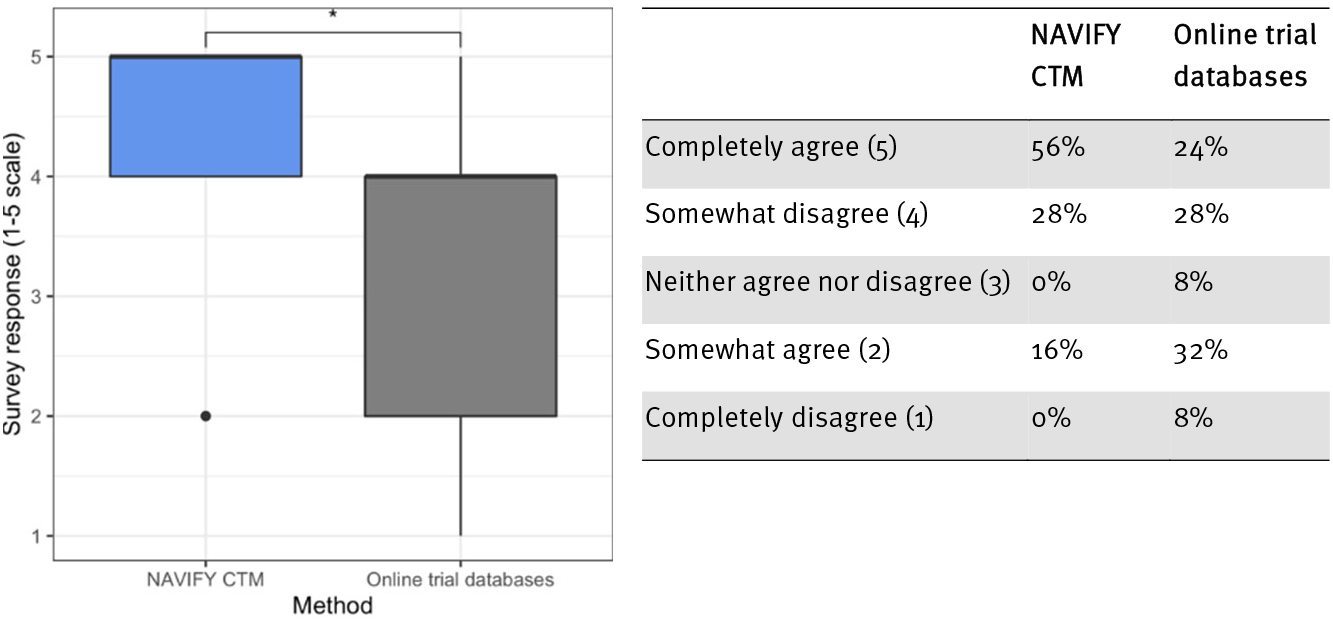
Participants’ perception of having enough time to complete the trial matching exercise (by method). Participants reported a significantly higher degree of agreement with having sufficient time to complete the trial matching exercise using NAVIFY CTM (p = 0.020). Asterisk (*) indicates results of statistical significance.

#### Quality of trial matches

There was no statistically significant difference in the quality of trial matches between NAVIFY CTM and all other online trial databases combined (*p* = 0.790). There was a statistically significant difference when comparing NAVIFY CTM to NWL CRN Lung Cancer Portfolio (*p* = 0.026) and NIHR Be Part of Research (*p* = 0.040), although there were few trial matches using these databases (*n* = 21 and *n* = 8, respectively). There were no statistically significant differences compared with ClinicalTrials.gov (*p* = 0.590), the EU Clinical Trials Register (*p* = 0.840), or NIHR CRN Portfolio Search (*p* = 0.690) (Figure 3).

**Figure 3.**
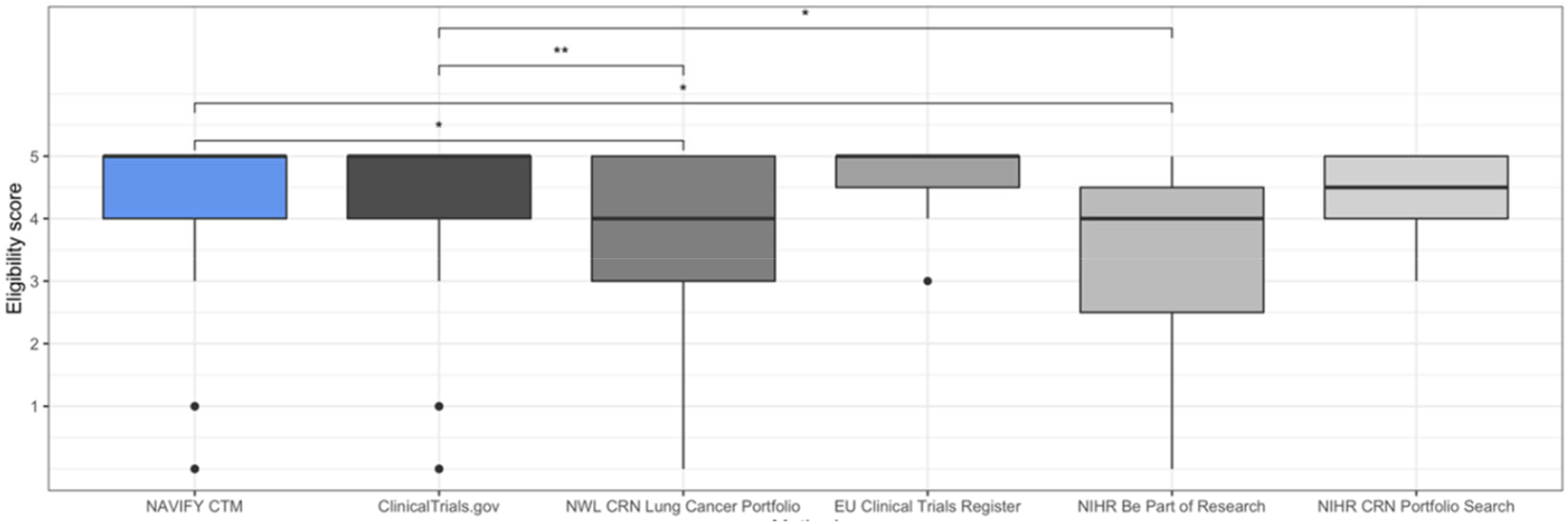
Comparison of eligibility scores for trial matches using NAVIFY CTM and using online trial databases. Single asterisk (^*^) indicates results of statistical significance, double asterisk (^**^) indicates non-significant results.

On the post-simulation survey, participants were more likely to agree that trial suggestions via NAVIFY CTM and ClinicalTrials.gov were relevant compared with other online trial databases. No statistically significant differences were identified between NAVIFY CTM and the next best scoring databases, ClinicalTrials.gov (*p* = 0.780), the EU Clinical Trials Register (*p* = 0.150), and NIHR Be Part of Research (*p* = 0.150). Both NAVIFY CTM and ClinicalTrials.gov scored significantly higher compared to NIHR CRN Portfolio Research (*p* = 0.049 and *p* = 0.049, respectively), and ISRCTN (*p* = 0.049 and *p* = 0.008, respectively) (Figure 4).

**Figure 3.**
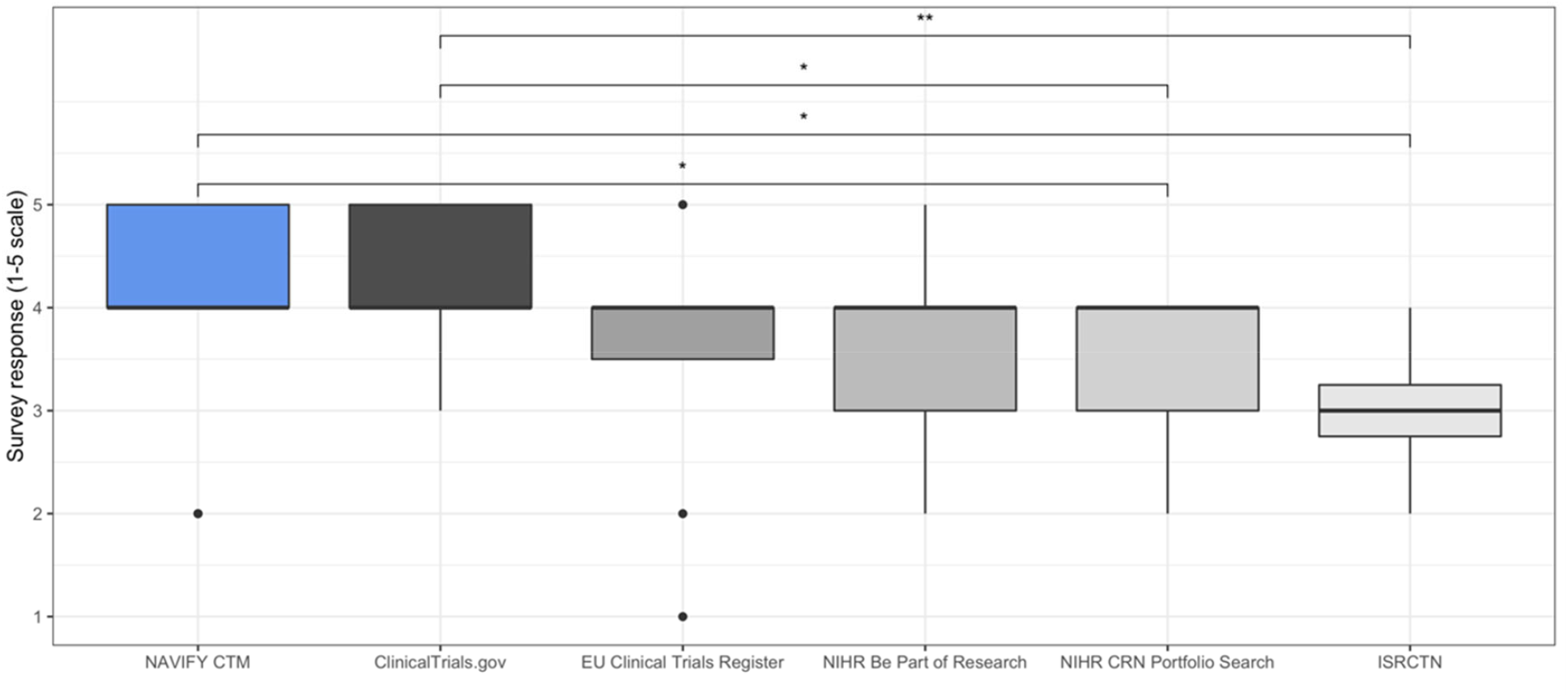
Perceived relevance of top trial suggestions as per post-simulation survey (NAVIFY CTM vs. online trial databases). Participants were significantly more likely to agree that trial suggestions via NAVIFY CTM and ClinicalTrials.gov were relevant compared with the other online trial databases. Single asterisk (^*^) indicates results of statistical significance, double asterisk (^**^) indicates non-significant results.

#### Cognitive burden

Participants were significantly more likely to agree that less mental effort was required to complete trial matching using NAVIFY CTM compared to all other online trial databases combined (*p* = 0.001). The difference was significant when compared individually with each of the online trial databases, including ClinicalTrials.gov (*p* = 0.002), the EU Clinical Trials Register (*p* = 0.001), NIHR Be Part of Research (*p* = 0.047), NIHR CRN Portfolio Search (*p* = 0.005), and the ISRCTN (*p* = 0.039). Participants were more likely to require ‘very low’, ‘low’ or ‘rather low’ mental effort to use NAVIFY CTM (*n* = 11, 44%) compared to all online trial databases taken together (*n* = 2, 8%). This difference remained when compared against online trial databases individually (Figure 5) (Table 4).

**Table 4.** Measure of mental effort required to use NAVIFY CTM compared to online trial databases. Summary table of survey responses via the Paas Scale

|  | NAVIFY CTM | ClinicalTrials.gov | EU Clinical Trials Register | NIHR Be Part of Research | NIHR CRN Portfolio Search | ISRCTN |
| --- | --- | --- | --- | --- | --- | --- |
| 9 = very, very high mental effort | 0% | 4% | 13% | 0% | 13% | 50% |
| 8 = very high mental effort | 0% | 8% | 25% | 14% | 0% | 0% |
| 7 = rather high mental effort | 20% | 56% | 38% | 29% | 50% | 0% |
| 6 = high mental effort | 12% | 8% | 13% | 14% | 25% | 25% |
| 5 = neither low nor high mental effort | 24% | 8% | 13% | 43% | 13% | 25% |
| 4 = low mental effort | 12% | 0% | 0% | 0% | 0% | 0% |
| 3 = rather low mental effort | 28% | 12% | 0% | 0% | 0% | 0% |
| 2 = very low mental effort | 4% | 4% | 0% | 0% | 0% | 0% |
| 1 = very, very low mental effort | 0% | 0% | 0% | 0% | 0% | 0% |

**Figure 5.**
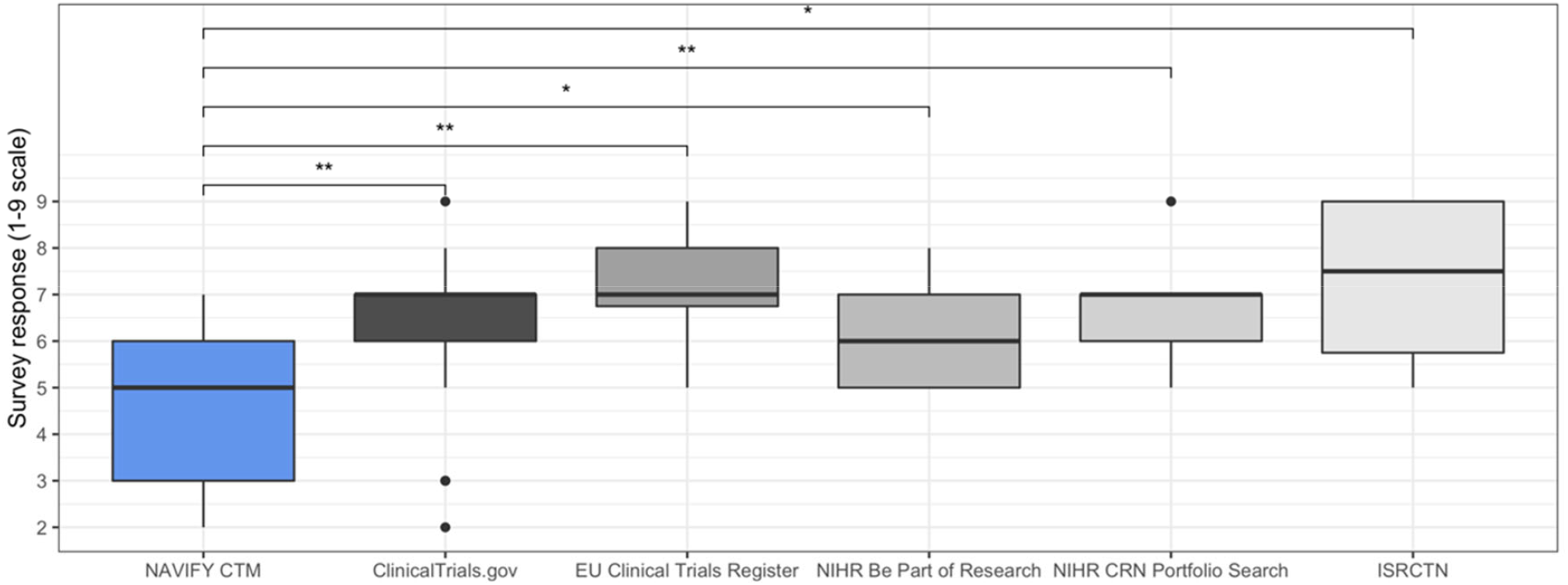
Measure of mental effort required to use NAVIFY CTM compared to online trial databases. Participants reported that less mental that less mental effort was required to use NAVIFY CTM and this difference was statistically significant compare to all online trial databases (p = 0.001). Single asterisk (*) indicates results of statistical significance, double asterisk (^**^) indicates non-significant results.

Cognitive burden was measured objectively using the Stroop Color and Word Test (SCWT). The Stroop effect – defined as the difference in average reaction times (RTs) for incongruent and congruent Stoop tasks – was measured before and after each trial matching exercise. The Stroop effect after using NAVIFY CTM was equal to the Stroop effect after using online trial databases. The difference in average RT before and after using NAVIFY CTM was smaller (28 msec) than the same difference for online trial databases (36 msecs), although the difference was not statistically significant (*p* = 0.662 and *p* = 0.776, respectively). Stroop task errors were higher before trial matching using online trial databases compared to after (3.1% vs. 2.4%). For NAVIFY CTM, errors were lower before trial matching compared to after (2.3% vs 2.6%). This difference in errors before and after each trial matching exercise was not statistically significant (*p* = 0.993 and *p* = 0.092, respectively).

## Discussion

### Summary and interpretation of key findings

The hypothesis of the study was that participants would identify trial matches quicker using NAVIFY CTM than online trial databases. While the difference in time-to-match between the tool approaches was found to be marginally non-significant (*p* = 0.063), this suggests that we could demonstrate a statistically significant difference with a larger sample size. Interestingly, participants were significantly more likely to feel like they had enough time to complete the exercise when using NAVIFY CTM (*p* = 0.020). This effect could be explained by the lower mental effort required to use NAVIFY CTM or as a result of bias towards the solution being evaluated due to the absence of blinding. Risk of bias could be reduced using a single- or double-blind study design, blinding of any kind can be difficult when evaluating a novel digital health solution. Alternatively, a between-subject study design could be used to mitigate bias amongst participants.

There was no statistically significant difference in the quality of trial matches between NAVIFY CTM and all other online trial databases combined (p = 0.790). However, participants perceived that the trial match suggestions from NAVIFY CTM and ClinicalTrials.gov were more relevant compared to the other online trial databases. This finding could be due to factors affecting user satisfaction which caused participants to favor NAVIFY CTM, such as information format, content, consistency and ease of navigation^56^. However, this effect could also be due to information biases which effect participants’ judgement, i.e. participants may have been influenced or biased towards providing positive feedback about trial matches from NAVIFY CTM because they knew the tool was developed to provide better matches than other online trial databases^57^.

This study objectively and subjectively measured the cognitive burden of participants associated with clinical tasks, using the Stroop Color Word Test (SCWT) and Paas scale respectively. SCWT is one of the most widely used paradigms and has been applied in the clinical setting^58–62^. While no difference could be ascertained on the SCWT, analysis of Paas scale responses showed that there was lower perceived mental effort with using NAVIFY CTM. The challenges associated with decision-making in healthcare contexts are well understood and associated with high levels of cognitive load^40^. Therefore, there is a need for solutions that reduce the cognitive load associated with decision-making processes, including clinical trial matching. However, there is limited research on sensitive and objective measurement instruments for the cognitive burden of a task placed on healthcare professionals. Behavior paradigms to measure executive ability or mental agility have been well established in the Experimental Psychology and Neuroscience^58,63,64^. Future work could explore the use of other paradigms to measure the brain’s response to a cognitive event, such as the event-related potential (ERP) paradigm^65^.

Overall, most participants agreed that they had sufficient guidance to participate in the simulations (n = 24, 96%) and that the synthetic patient cases were detailed enough (n = 18, 72%). According to participants the main benefits of using SBR to evaluate digital health solutions are that it creates an opportunity for clinicians to preview the solution and provide feedback before it is implemented in clinical practice. The main perceived challenges were the additional amount of time required to learn the new tool and not having enough time to complete the tasks in the simulation.

### Comparison with previous literature

Prior to this study, we piloted our methodology by conducting simulation sessions with 10 clinicians, five with experience matching patients to cancer clinical trials in the UK (experts) and five without experience (non-experts). The objective of the pilot phase of the study was to rapidly test and refine the SBR methods (including data collection procedures) with a smaller number of participants. The results of the pilot phase are reported in a separate abstract published by the research team [in submission].

A comparison of the study methods for the pilot and study phase is shown in Table 5. NAVIFY CTM was updated between phases (from v1.3 to v1.4). This allowed us to assess two different versions of the software in quick succession. Changes were also made to the SCWT between the pilot and study phases. Based on the review of studies where the time limit is not calibrated^66,67^, a time limit of 2.0 seconds was set for all participants in the pilot. We observed that the accuracy across all participants was above 97%, suggesting a ceiling effect (i.e., performance was not significantly impacted by mental agility). The time limit was therefore shortened to 1.5 seconds for the study phase, however, accuracy rates remained above 95%, suggesting that the ceiling effect remained. In future studies, reaction times could be further shortened, or individual calibration procedures applied. Second, to reduce learning effect, a single practice SCWT consisting of 10 Stroop tasks was introduced before the first full SCWT.

**Table 5.** Comparison of study design and data collection procedures between the pilot and study phases.

|  | **Pilot phase** | **Study phase\*** |
| --- | --- | --- |
| Primary objective(s) | Rapidly test methodology and data collection | Evaluate efficiency, quality, and cognitive burden of trial matching |
| Sample size | $n = 10$ (5 experts, 5 non-experts) | $n = 25$ (all experts) |
| NAVIFY CTM software version | v1.3 | v1.4 |
| **Comparator** |  |  |
| Presentation of patient cases | Printed materials | Digital (PDF and JPG files) |
| Online trial databases | ClinicalTrials.gov | ClinicalTrials.gov |
|  | EU Clinical Trials Register | EU Clinical Trials Register |
|  | NIHR Be Part of Research | NIHR Be Part of Research |
|  | NIHR CRN Portfolio Search | NIHR CRN Portfolio Search |
|  | ISRCTN | ISRCTN |
|  |  | NWL CRN Lung Cancer Portfolio |
| **Data collection** |  |  |
| Efficiency | Time to complete each trial match | Time to complete each trial match |
| Quality | Eligibility score assigned by thoracic oncologist | Eligibility score assigned by thoracic oncologist |
| Cognitive burden | SCWT<br>No practice test<br>Time limit per task = 2 seconds<br>Paas Scale | SCWT<br>Practice test = 10 tasks<br>Time limit per task = 1.5 seconds<br>Paas Scale |
| Use of SBR | Post-simulation survey | Post-simulation survey |

\* The Results section of this article reports those from the study phase alone.

### Strengths and limitations

This study employed a within-subject design in which the evaluation of NAVIFY CTM and online trial databases were performed in the same simulated environment by the same participants. The ability to reproduce the same simulated clinical environment before and after, and for all participants, increases the confidence that any measurable differences between NAVIFY CTM and online trial databases can be attributed to the tools themselves as opposed to external factors^18^.

This study also demonstrated the value of using synthetic patient data, allowing for the timely evaluation of NAVIFY CTM whilst maintaining the fidelity of the simulated clinical scenario. Using real patient data in research introduces additional ethical and information governance considerations, such as data privacy and confidentiality. It typically involves de-identifying or pseudonymizing data, adding “noise” to data or grouping variables together. However, these approaches are subject to risk as well as delays obtaining data^68,69^. Using synthetic patient data in this study mitigated the risks associated with using de-identified or pseudonymized patient data as well as the need to undergo ethics and data security approval to obtain real patient data. The key benefit of this approach is that the duration of study set-up was relatively quick compared to studies which require additional permissions in place because of the use of real patient data^8^. This demonstrates the value of using synthetic patient data to conduct evaluations of digital health solutions in a timeframe that is appropriate for the fast-paced nature of innovation and product development.

To ensure that the synthetic patient cases could be used to evaluate the ability of NAVIFY CTM to identify appropriate trial matches they were developed to match to the eligibility criteria of at least one actively enrolling cancer clinical trial in London. Furthermore, the level of clinical detail had to be reflective of what clinicians would encounter in routine practice to ensure the simulated clinical scenario felt realistic to participants. To ensure the fidelity of the synthetic patient cases, they were developed by practicing consultants with extensive experience of lung cancer patient cases in their routine practice. By instructing the consultants to develop the cases with the level of detail and the type of format in which patient data is usually presented this ensured the synthetic patient cases were realistic to research participants and sufficient in detail to evaluate the effectiveness of NAVIFY CTM.

One challenge of the study was identifying an appropriate method to determine the quality or relevance of the trial matches identified by participants. Although the eligibility criteria of clinical trials are a conclusive measure of a patient’s suitability, the process is currently governed by the subjective approach taken by individual clinicians and research staff members. In this study, engaging a consultant thoracic oncologist to independently score trial matches according to eligibility criteria was an effective proxy for measuring the quality of trial matches. In this way, distinctions could be made between trial matches in which all of the eligibility criteria were met or partially met.

The main benefit of SBR is the ability to collect measurements of participants’ responses during the study and simulated scenarios in ways that would be otherwise impossible or impractical in clinical settings such as pilot programs or observational studies. In this study, we were able to obtain extensive measurements related to cognitive burden with the Stroop Color Word Test (SCWT), record screen activity and gather additional feedback from participants immediately after they conducted the trial matching exercises. It would not be practical or in the best interest of patient care to gather such measurements and feedback during a real-world evaluation conducted in a clinical setting. Therefore, the use of SBR allowed for granular evaluations of participant response and real-time feedback of NAVIFY CTM which could support product development and refinement before it is implemented in routine clinical practice.

For this research study, ethics approval was obtained via the university’s Research Governance and Integrity Team, a process that typically takes two to three weeks. Approval from the Health Research Authority in England – with a typical timeline of two to three months – was not required as the study did not involve real patients or real patient data, did not take place at an NHS organization, and participants were not recruited via the NHS.

This study employed a novel methodology to evaluate a digital health solution with practicing clinicians and research staff outside of a clinical setting, therefore there are some limitations and areas for improvement.

This study was conducted during the COVID-19 pandemic and lockdown measures were in place which impacted travel^70,71^, as well as the availability of clinicians to participate in research. This may have created a bias towards participants who were located closer to St Mary’s Hospital, London or who had more flexible clinical commitments. Furthermore, participants were recruited to complete trial matching exercises individually, while the literature on cancer clinical trial recruitment in the UK shows some evidence of multidisciplinary team involvement in decision-making^37,72^.

It is best practice to calibrate reaction time limit for each participant and to control for sleep and caffeine intake when using the SCWT in experimental psychology^42^. To calibrate the optimal reaction time limit, the time limit is set at the point where each participant reaches 80% accuracy, and typically takes 30 minutes per participant. Due to the clinical responsibilities of study participants and the resulting time constraints for conducting the sessions, it was not reasonable to impose these controls.

One of the limitations of SBR methods is the challenge of assessing how realistic the simulated events and synthetic data are compared to routine clinical settings and real patient data. Although most participants reported that the synthetic patient cases were realistic, the use of real patient data in the study could have yielded other insights from participants regarding the usability and applicability of the tool in clinical practice. Future work in this area could explore alternative approaches to develop more detailed synthetic patient cases and synthetic electronic health record data, which reflect the presentation and granularity of information seen in routine practice^68,69,73^.

## Implications for practice and future opportunities

This study demonstrates the ability to evaluate a novel clinical decision support tool in a simulated clinical scenario using synthetic patient cases. We were able to collect quantitative insights on the performance of NAVIFY CTM compared to online trial databases and to collect valuable user feedback without the need for implementation in clinical practice.

The COVID-19 pandemic has had widespread consequences on public and healthcare professionals’ attitudes towards digital health solutions and resulted in the expedition of policies to support the adoption of digital health solutions and digital-first models of care^74–77^. However, appropriate and robust methods for evaluating digital health solutions have been slow to materialize, in particular approaches that reflect the iterative and fast-paced nature of innovation. This study demonstrates the ability to gather early insights on the performance of a digital health solution and the opportunities for implementation in clinical practice. The use of SBR also seems sufficient to derive user experience insights from practicing clinicians regarding the usability and acceptability of clinical decision support tools. These findings justify the use of SBR methodology to generate insights that support product development of low-risk digital health solutions before they are implemented in clinical practice.

Another future area of research could be the use of SBR to conduct remote evaluations of digital health solutions. This has important implications in light of the COVID-19 pandemic, which has resulted in shifts to remote working. Guidance published by Public Health England has supported the use of remote approaches to evaluate digital health solutions, as opposed to face-to-face approaches^78^. For certain digital health solutions, such as clinical workflow solutions, teleconsultation services or any other software-based solutions, the design and methodology used in this study could be translated to remote settings and enable evaluations in different geographical locations simultaneously. This approach could generate evidence within a short timeframe, further enhancing the potential for return on investment for innovators.

## Data Availability

All data generated or analysed during this study are included in this published article (and its supplementary information files).

## Acknowledgements

None

## References

1. Lefevre A, Agarwal S, Zeller K, Vasudevan L, Healey K, Tamrat T, et al. Monitoring and evaluating digital health interventions: a practical guide to conducting research and assessment. 2016.

2. Snowdon DA. Digital Health: A Framework For Healthcare Transformation. HIMSS; p. 70.

3. Evidence standards framework for digital health technologies | Our programmes | What we do | About [Internet]. NICE. NICE; [cited 2020 Oct 30]. Available from: https://www.nice.org.uk/about/what-we-do/our-programmes/evidence-standards-framework-for-digital-health-technologies

4. Why does the NHS struggle to adopt eHealth innovations? A review of macro, meso and micro factors | BMC Health Services Research | Full Text [Internet]. [cited 2020 Oct 30]. Available from: https://bmchealthservres.biomedcentral.com/articles/10.1186/s12913-019-4790-x

5. Shaping the future of UK healthcare [Internet]. Deloitte United Kingdom. [cited 2021 Feb 3]. Available from: https://www2.deloitte.com/uk/en/pages/life-sciences-and-healthcare/articles/shaping-the-future-of-uk-healthcare.html

6. Montgomery HE, Haines A, Marlow N, Pearson G, Mythen MG, Grocott MPW, et al. The future of UK healthcare: problems and potential solutions to a system in crisis. Ann Oncol. 2017 Aug 1;28(8):1751–5.

7. De Silva D. What’s getting in the way? [Internet]. The Health Foundation; 2015 Feb [cited 2020 Oct 30]. Available from: https://reader.health.org.uk/whats-getting-in-the-way

8. Guo C, Ashrafian H, Ghafur S, Fontana G, Gardner C, Prime M. Challenges for the evaluation of digital health solutions—A call for innovative evidence generation approaches. Npj Digit Med. 2020 Aug 27;3(1):1–14.

9. Mathews SC, McShea MJ, Hanley CL, Ravitz A, Labrique AB, Cohen AB. Digital health: a path to validation. Npj Digit Med. 2019 May 13;2(1):1–9.

10. Hollis C, Falconer CJ, Martin JL, Whittington C, Stockton S, Glazebrook C, et al. Annual Research Review: Digital health interventions for children and young people with mental health problems – a systematic and meta-review. J Child Psychol Psychiatry. 2017;58(4):474–503.

11. Singh K, Drouin K, Newmark LP, Lee J, Faxvaag A, Rozenblum R, et al. Many Mobile Health Apps Target High-Need, High-Cost Populations, But Gaps Remain. Health Aff Proj Hope. 2016 Dec 1;35(12):2310–8.

12. Murray E, Hekler EB, Andersson G, Collins LM, Doherty A, Hollis C, et al. Evaluating Digital Health Interventions: Key Questions and Approaches. Am J Prev Med. 2016 Nov 1;51(5):843–51.

13. O’Connor S, Hanlon P, O’Donnell CA, Garcia S, Glanville J, Mair FS. Understanding factors affecting patient and public engagement and recruitment to digital health interventions: a systematic review of qualitative studies. BMC Med Inform Decis Mak. 2016 Sep 15;16(1):120.

14. O’Connor S, Hanlon P, O’Donnell CA, Garcia S, Glanville J, Mair FS. Barriers and facilitators to patient and public engagement and recruitment to digital health interventions: protocol of a systematic review of qualitative studies. BMJ Open. 2016 Sep 1;6(9):e010895.

15. Pratap A, Neto EC, Snyder P, Stepnowsky C, Elhadad N, Grant D, et al. Indicators of retention in remote digital health studies: a cross-study evaluation of 100,000 participants. Npj Digit Med. 2020 Feb 17;3(1):1–10.

16. Ong J, Parchment V, Zheng X. Effective regulation of digital health technologies. J R Soc Med. 2018 Dec 1;111(12):439–43.

17. Greaves F, Joshi I, Campbell M, Roberts S, Patel N, Powell J. What is an appropriate level of evidence for a digital health intervention? The Lancet. 2018 Dec 22;392(10165):2665–7.

18. Lamé G, Dixon-Woods M. Using clinical simulation to study how to improve quality and safety in healthcare. BMJ Simul Technol Enhanc Learn [Internet]. 2020 Mar 1 [cited 2021 Feb 3];6(2). Available from: https://stel.bmj.com/content/6/2/87

19. Aebersold M. Simulation-based learning: No longer a novelty in undergraduate education. Online J Issues Nurs. 2018 May 1;23:1–1.

20. Kim J, Park J-H, Shin S. Effectiveness of simulation-based nursing education depending on fidelity: a meta-analysis. BMC Med Educ [Internet]. 2016 May 23 [cited 2020 Oct 30];16. Available from: https://www.ncbi.nlm.nih.gov/pmc/articles/PMC4877810/

21. Higham H, Baxendale B. To err is human: use of simulation to enhance training and patient safety in anaesthesia. BJA Br J Anaesth. 2017 Dec 1;119(Suppl_1):i106–14.

22. Pottle J. Virtual reality and the transformation of medical education. Future Heal J. 2019 Oct 1;6(3):181–5.

23. Cook DA, Brydges R, Zendejas B, Hamstra SJ, Hatala R. Mastery Learning for Health Professionals Using Technology-Enhanced Simulation: A Systematic Review and Meta-Analysis. Acad Med. 2013 Aug;88(8):1178–86.

24. Gaba DM. The Future Vision of Simulation in Healthcare. Simul Healthc. 2007 Jul;2(2):126–35.

25. Marshall SD, Flanagan B. Simulation-based education for building clinical teams. J Emerg Trauma Shock. 2010;3(4):360–8.

26. Martin A, Cross S, Attoe C. The Use of in situ Simulation in Healthcare Education: Current Perspectives. Adv Med Educ Pract. 2020 Nov 27;11:893–903.

27. Freund D, Andersen PO, Svane C, Meyhoff CS, Sørensen JL. Unannounced vs announced in situ simulation of emergency teams: Feasibility and staff perception of stress and learning. Acta Anaesthesiol Scand. 2019;63(5):684–92.

28. Sørensen JL, Lottrup P, Vleuten C van der, Andersen KS, Simonsen M, Emmersen P, et al. Unannounced in situ simulation of obstetric emergencies: staff perceptions and organisational impact. Postgrad Med J. 2014 Nov 1;90(1069):622–9.

29. Sørensen JL, Østergaard D, LeBlanc V, Ottesen B, Konge L, Dieckmann P, et al. Design of simulation-based medical education and advantages and disadvantages of in situ simulation versus off-site simulation. BMC Med Educ. 2017 Jan 21;17(1):20.

30. Katsaliaki K, Mustafee N. Applications of simulation within the healthcare context. J Oper Res Soc. 2011 Aug 1;62(8):1431–51.

31. Cooper K, Brailsford SC, Davies R. Choice of modelling technique for evaluating health care interventions. J Oper Res Soc. 2007 Feb 1;58(2):168–76.

32. Low D, Clark N, Soar J, Padkin A, Stoneham A, Perkins GD, et al. A randomised control trial to determine if use of the iResus© application on a smart phone improves the performance of an advanced life support provider in a simulated medical emergency. Anaesthesia. 2011 Apr;66(4):255–62.

33. Jensen S, Kushniruk AW, Nøhr C. Clinical simulation: A method for development and evaluation of clinical information systems. J Biomed Inform. 2015 Apr 1;54:65–76.

34. Ammenwerth E, Hackl WO, Binzer K, Christoffersen TEH, Jensen S, Lawton K, et al. Simulation Studies for the Evaluation of Health Information Technologies: Experiences and Results. Health Inf Manag J. 2012 Jun 1;41(2):14–21.

35. The Role of Clinical Trial Participation in Cancer Research: Barriers, Evidence, and Strategies | American Society of Clinical Oncology Educational Book [Internet]. [cited 2020 Dec 7]. Available from: https://ascopubs.org/doi/full/10.1200/EDBK_156686

36. Record number of patients take part in clinical research [Internet]. [cited 2020 Jun 16]. Available from: https://www.nihr.ac.uk/news/record-number-of-patients-take-part-in-clinical-research/11746?diaryentryid=44785

37. Cox K, Avis M, Wilson E, Elkan R. An evaluation of the introduction of Clinical Trial Officer roles into the cancer clinical trial setting in the UK. Eur J Cancer Care (Engl). 2005;14(5):448–56.

38. Wilson C, Rooshenas L, Paramasivan S, Elliott D, Jepson M, Strong S, et al. Development of a framework to improve the process of recruitment to randomised controlled trials (RCTs): the SEAR (Screened, Eligible, Approached, Randomised) framework. Trials. 2018 Dec;19(1):1–10.

39. Patel VL, Kaufman DR, Arocha JF. Emerging paradigms of cognition in medical decision-making. J Biomed Inform. 2002 Feb 1;35(1):52–75.

40. Burgess DJ. Are Providers More Likely to Contribute to Healthcare Disparities Under High Levels of Cognitive Load? How Features of the Healthcare Setting May Lead to Biases in Medical Decision Making. Med Decis Making. 2010 Mar 1;30(2):246–57.

41. Goodman LA. Snowball Sampling. Ann Math Stat. 1961;32(1):148–70.

42. Tanaka M, Ishii A, Shigihara Y, Tajima S, Funakura M, Kanai E, et al. Impaired Selective Attention Caused By Mental Fatigue. J Neurol Sci. 2012 Jan 1;29:542–53.

43. Stoet G. PsyToolkit: a software package for programming psychological experiments using Linux. Behav Res Methods. 2010 Nov;42(4):1096–104.

44. Stoet G. PsyToolkit: A Novel Web-Based Method for Running Online Questionnaires and Reaction-Time Experiments. Teach Psychol. 2017 Jan 1;44(1):24–31.

45. ClinicalTrials.gov [Internet]. [cited 2021 May 4]. Available from: https://clinicaltrials.gov/

46. EU Clinical Trials Register [Internet]. [cited 2021 May 4]. Available from: https://www.clinicaltrialsregister.eu/

47. NIHR CRN Portfolio Search [Internet]. [cited 2021 May 4]. Available from: https://public-odp.nihr.ac.uk/QvAJAXZfc/opendoc.htm?document=crncc_users%5Cfind%20a%20clinical%20research%20study.qvw&lang=en-US&host=QVS%40crn-prod-odp-pu&anonymous=true

48. Be Part of Research [Internet]. [cited 2020 Jun 16]. Available from: https://bepartofresearch.nihr.ac.uk/

49. ISRCTN Registry [Internet]. [cited 2021 May 4]. Available from: https://www.isrctn.com/

50. CRN North West London [Internet]. [cited 2021 May 4]. Available from: https://local.nihr.ac.uk/lcrn/north-west-london/

51. Paas FGWC, Van Merriënboer JJG. The Efficiency of Instructional Conditions: An Approach to Combine Mental Effort and Performance Measures. Hum Factors. 1993 Dec 1;35(4):737–43.

52. Sweller J. Measuring cognitive load. Perspect Med Educ. 2018 Feb 1;7(1):1–2.

53. Braun V, Clarke V. Using thematic analysis in psychology. Qual Res Psychol. 2006 Jan 1;3(2):77–101.

54. R Core Team. R: A language and environment for statistical computing. R Foundation for Statistical Computing [Internet]. Vienna, Austria; 2020. Available from: https://www.R-project.org/

55. Millard SP. EnvStats, an R Package for Environmental Statistics. In: Wiley StatsRef: Statistics Reference Online [Internet]. American Cancer Society; 2014 [cited 2021 Mar 18]. Available from: https://onlinelibrary.wiley.com/doi/abs/10.1002/9781118445112.stat07181

56. Sindhuja PN, Dastidar SG. Impact of the Factors Influencing Website Usability on User Satisfaction. IUP J Manag Res. 2009 Dec;8(12):54–66.

57. Hartmann J, De Angeli A, Sutcliffe A. Framing the user experience: information biases on website quality judgement. In: Proceedings of the SIGCHI Conference on Human Factors in Computing Systems [Internet]. New York, NY, USA: Association for Computing Machinery; 2008 [cited 2021 May 4]. p. 855–64. (CHI ‘08). Available from: https://doi.org/10.1145/1357054.1357190

58. Luck SJ, Gold JM. The Translation of Cognitive Paradigms for Patient Research. Schizophr Bull. 2008 Jul 1;34(4):629–44.

59. Guarino A, Favieri F, Boncompagni I, Agostini F, Cantone M, Casagrande M. Executive Functions in Alzheimer Disease: A Systematic Review. Front Aging Neurosci [Internet]. 2019 [cited 2021 Mar 23];10. Available from: https://www.frontiersin.org/articles/10.3389/fnagi.2018.00437/full

60. Epp AM, Dobson KS, Dozois DJA, Frewen PA. A systematic meta-analysis of the Stroop task in depression. Clin Psychol Rev. 2012 Jun 1;32(4):316–28.

61. Jacobs JV, Kasser SL. Effects of dual tasking on the postural performance of people with and without multiple sclerosis: a pilot study. J Neurol. 2012 Jun 1;259(6):1166–76.

62. Denney DR, Lynch SG. The impact of multiple sclerosis on patients’ performance on the Stroop Test: processing speed versus interference. J Int Neuropsychol Soc JINS. 2009 May;15(3):451–8.

63. Barch DM, Carter CS, Arnsten A, Buchanan RW, Cohen JD, Geyer M, et al. Selecting Paradigms From Cognitive Neuroscience for Translation into Use in Clinical Trials: Proceedings of the Third CNTRICS Meeting. Schizophr Bull. 2009 Jan 1;35(1):109–14.

64. Kessels RPC. Improving precision in neuropsychological assessment: Bridging the gap between classic paper-and-pencil tests and paradigms from cognitive neuroscience. Clin Neuropsychol. 2019 Feb 17;33(2):357–68.

65. Ghani U, Signal N, Niazi IK, Taylor D. A novel approach to validate the efficacy of single task ERP paradigms to measure cognitive workload. Int J Psychophysiol. 2020 Dec 1;158:9–15.

66. Jung M, Jonides J, Berman MG, Northouse L, Koelling TM, Pressler SJ. Construct Validity of the Multi-Source Interference Task to Examine Attention in Heart Failure. Nurs Res. 2018 Dec;67(6):465–72.

67. Kandul S, Naguleswaran A. Lying under self-control depletion and time pressure [Internet]. IRENE Working Paper; 2019 [cited 2021 Mar 18]. Report No.: 19–05. Available from: https://www.econstor.eu/handle/10419/213480

68. Goncalves A, Ray P, Soper B, Stevens J, Coyle L, Sales AP. Generation and evaluation of synthetic patient data. BMC Med Res Methodol. 2020 May 7;20(1):108.

69. Tucker A, Wang Z, Rotalinti Y, Myles P. Generating high-fidelity synthetic patient data for assessing machine learning healthcare software. Npj Digit Med. 2020 Nov 9;3(1):1–13.

70. London, South Essex, and South Hertfordshire to move to Tier 3 restrictions [Internet]. GOV.UK. [cited 2021 Feb 23]. Available from: https://www.gov.uk/government/news/london-south-essex-and-south-hertfordshire-to-move-to-tier-3-restrictions

71. Prime Minister announces Tier 4: ‘Stay At Home’ Alert Level in response to new COVID variant [Internet]. GOV.UK. [cited 2021 Feb 23]. Available from: https://www.gov.uk/government/news/prime-minister-announces-tier-4-stay-at-home-alert-level-in-response-to-new-covid-variant

72. Maslin-Prothero S. The role of the multidisciplinary team in recruiting to cancer clinical trials. Eur J Cancer Care (Engl). 2006;15(2):146–54.

73. Chen J, Chun D, Patel M, Chiang E, James J. The validity of synthetic clinical data: a validation study of a leading synthetic data generator (Synthea) using clinical quality measures. BMC Med Inform Decis Mak. 2019 Mar 14;19(1):44.

74. Kadakia K, Patel B, Shah A. Advancing digital health: FDA innovation during COVID-19. Npj Digit Med. 2020 Dec 17;3(1):1–3.

75. Rights (OCR) O for C. Notification of Enforcement Discretion for Telehealth [Internet]. HHS.gov. 2020 [cited 2021 Feb 23]. Available from: https://www.hhs.gov/hipaa/for-professionals/special-topics/emergency-preparedness/notification-enforcement-discretion-telehealth/index.html

76. Teles M, Sacchetta T, Matsumoto Y. COVID-19 Pandemic Triggers Telemedicine Regulation and Intensifies Diabetes Management Technology Adoption in Brazil. J Diabetes Sci Technol. 2020 Jul 1;14(4):797–8.

77. Gerke S, Stern AD, Minssen T. Germany’s digital health reforms in the COVID-19 era: lessons and opportunities for other countries. Npj Digit Med. 2020 Jul 10;3(1):1–6.

78. Rapid evaluation of digital health products during the COVID-19 pandemic [Internet]. GOV.UK. [cited 2021 Mar 8]. Available from: https://www.gov.uk/guidance/rapid-evaluation-of-digital-health-products-during-the-covid-19-pandemic

