## Supplementary Information for "Evaluation of a clinical decision support tool for matching cancer patients to clinical trials using simulation-based research"

### Simulation session guide

Imperial College London

#### NAVIFY Clinical Trial Match evaluation – simulation session guide

##### Background

This is an independent study conducted by the team at the Institute of Global Health Innovation.

The NAVIFY Clinical Trial Match application is a digital solution that helps clinicians to match patients to clinical trials. The app is part of the NAVIFY Tumor Board solution, a solution used to present patient information during cancer multidisciplinary team (MDT) meetings.

NAVIFY Clinical Trial Match automatically searches a patient record using key criteria to identify trial matches (e.g., cancer type, stage, genomic alterations, etc.). Further information on how the app works is provided separately in the NAVIFY Clinical Trial Match User Guide.

At this stage of the product development, Roche would like to understand how NAVIFY Clinical Trial Match app could be implemented in UK healthcare settings and where it could be useful in clinical practice.

SAMPLE

##### Purpose of today

The purpose of today is to evaluate the Clinical Trial Match app, including how it could improve the practice of screening patients for clinical trials, how relevant the search results are, and how easy it is to use compared to standard methods and tools.

##### Simulated Scenario

In this session, **you will play a member of the clinical or research team at Imperial College Healthcare NHS Trust who is responsible for screening patients with lung cancer for clinical trials** (e.g., Oncologist, Clinical Research Fellow, Research Nurse, Clinical Trial Practitioner, etc.).

Today you are preparing to meet with 10 patients in clinic and **you have been tasked with finding a suitable, actively recruiting clinical trial (or trials)** for each of them.

After a quick check, you and your team have already decided that **none of the patients are eligible for your local portfolio trials** and so you are searching for other trials nearby that might be suitable.

You will complete **two separate trial matching exercises today** (the order will be randomised):

1. For five of these patients, you will only have access to the NAVIFY Clinical Trial Match app to identify suitable trials.
2. For the five other patients, you will have access to other online search tools (ClinicalTrials.gov, ClinicalTrialsRegister.eu, NIHR's Portfolio Search website).

Imperial College London

**You will have exactly 60 minutes for each exercise.** During the trial matching exercise, you will be asked to record the suitable trials that you find on a worksheet.

#### Important assumptions for today's session

Regarding the patient cases:

- **Please try and find the trial(s) you think would be most suitable for each patient given the clinical information at hand.** There may be some investigations that have not been performed to make a final decision on enrolment. However, we would like you to focus on finding a potentially suitable trial rather than make any final decision on enrolment.
- **Please assume that all patient information is accurate.** The mock clinical details, pathology reports and radiology reports have been developed by experienced consultant oncologists, a histopathologist and a radiologist respectively.
- **Please consider the imaging indicative of what you would have to hand in a real-life situation.** For pragmatic reasons, the radiology imaging is in JPEG format (as opposed to full CT/MRI imaging).

Regarding NAVIFY:

- Please assume that NAVIFY Clinical Trial Match is already integrated with the electronic health records (EHR) system in your practice.
- Please assume that NAVIFY Clinical Trial Match is compliant with data protection, security and information governance requirements.

Regarding the simulation session:

- Throughout the session, the screen of the computer will be recorded to inform the research team's analysis. The recording will not be shared with anyone outside of the research team at Imperial College London.

SAMPLE

#### Session outcome

We hope to gather your anonymous feedback regarding:

- How useful you think NAVIFY Clinical Trial Match app would be in your routine practice
- Where you think NAVIFY Clinical Trial Match app could make a difference in the screening methods and decision-making processes in UK healthcare settings
- The simulation methods we are using today to evaluate this software, including the patient cases, the guidance materials provided, and the general organisation and conduct of the session.

Guidance provided to participants before the start of the simulation session which outlines the clinical tasks to be simulated using the study materials provided, as well as assumptions to made throughout the session regarding the patient cases and NAVIFY CTM.

### Example of a synthetic patient case including clinical summary, histopathology and radiology report

Imperial College Healthcare NHS Trust*

**CLINICAL DETAILS**

**BLOGGS, JOSEPH**
**IGHI005**
**DoB 1/1/1956**

64 yo Male

Diagnosis
October 2019, stage IV (T3N2M1a; T3N2M1a (separate nodules) lung adenoCa

November 2019 1L Pembrolizumab monotherapy initially q3w then q6w with good tolerance

Best response SD Feb 2020

2/6/2020 CT TAP: oligoprogressive disease in RUL
30/6/2020 CT TAP: new liver metastasis (12mm + 16mm) – further progression in RUL (radiology input)

PS ECOG 1
ASA 2

ex smoker 35 pack-years

Bloods: unremarkable but ALT 59 TSH 6.2 T4 12

SAMPLE

*this is a mock patient case prepared by a consultant oncologist for the purposes of the simulation sessions conducted by the Institute of Global Health Innovation, Imperial College London

Clinical summary of one of the synthetic patient cases which was composed by a consultant thoracic oncologist and reverse-engineered to match to an existing lung cancer clinical trial identified by the research team at Imperial College London.

NHS
Imperial College Healthcare
NHS Trust*

### HISTOPATHOLOGY REPORT

IGHI005/20
BLOGGS, JOSPEH

**SPECIMEN(S) RECEIVED**
EBUS, 4R
**CLINICAL DATA**
Previous RUL segmentectomy. Enlarging non FDG avid UR lymph node
**MACROSCOPIC DESCRIPTION**
Pot labelled BLOGGS, JOE and left 4R: Numerous tangled and brown and cream cores of tissue altogether measuring 14 x 12 x 2mm, all taken in 1 cassette. (CQ)
**MICROSCOPIC DESCRIPTION**
EBUS tissue comprising blood with fragments of crushed lymphoid tissue. There are clusters of atypical cells representing metastatic carcinoma.

Immunohistochemistry shows these to be positive for TTF1 and negative for p40. This represents metastatic lung adenocarcinoma.
**HISTOLOGICAL DIAGNOSIS**
EBUS TBNA 4R: metastatic adenocarcinoma
**COMMENT**
Report requires urgent review
***Electronically Signed Out*** Dr J Smith

SAMPLE

**SUPPLEMENTARY REPORT**
PD-L1 (22C3, DAKO Pharm DX) IHC Report
Sufficient for assessment – YES
100% of tumour cells expressing PD-L1
Intensity of staining by tumour cells:
3+ -80%
2+ - 20 %
1+ - 0 %
0 - 0 %
Staining Pattern: Homogenous expression
RESULT: STRONG POSITIVE (≥50% Expression)
***Electronically Signed Out*** Dr J Smith

**SUPPLEMENTARY REPORT**
ALK is negative by immunohistochemistry.
ROS1 is negative by immunohistochemistry.
***Electronically Signed Out*** Dr J Smith

*this is a mock patient case prepared by a consultant histopathologist for the purposes of the simulation sessions conducted by the Institute of Global Health Innovation, Imperial College London

NHS
Imperial College Healthcare
NHS Trust*

***Electronically Signed Out***    Dr J Smith

Detected Variants

TP53 p.(Pro72Arg), c.215C>G, COSM250061 45% TP53 p.(Asp281Asn), c.841G>A, COSM43596 21%

Detected Copy Number Gains

No copy number gains detected in this sample.

Detected Gene Fusions

No assessed gene fusion events were detected in this sample.

Salient 'negative' findings: Please be aware that due to the very high numbers of variants assessed by this assay, only detected variants are reported. Nevertheless, for the avoidance of doubt in relation to those genes of most common therapeutic interest; unless specifically stated to the contrary above, no variants were detected in this specimen at the following loci: BRAF (Codon 600), EGFR (Codons 492, 719, 768, 790, 797, 858, 861, exon 19 deletions/insertions or exon 20 insertions), KRAS & NRAS (Codons 12, 13, 59, 61, 117 & 146), KIT (exons 9, 11, 13 & 17), MET (transcribed sequences with exon 14 'skipped').

SAMPLE

*this is a mock patient case prepared by a consultant histopathologist for the purposes of the simulation sessions conducted by the Institute of Global Health Innovation, Imperial College London

Histology report for one of the synthetic patient cases which was developed by a consultant histopathologist and reverse-engineered to match to an existing lung cancer clinical trial identified by the research team at Imperial College London.

Imperial College Healthcare
NHS Trust*

### IMAGING REPORT

BLOGGS, JOSPEH

IGHI005

**RYJ20525263 02/06/2020**
CT Chest/Abdomen/Pelvis with contrast

Clinical details:
The 64 yrl male. Diagnosis of T3 N1 M1a lung adenoca. Nov 2019 treated with Pembrolizumab monotherapy initially q3w then q6w with good tolerance. Follow up scan.

Report:
Contrast enhanced axial images of the chest abdomen and pelvis.
Comparison is made with a previous CT scan of October 2019 and Feb 2020.

SAMPLE

Chest
The previous right upper lobe mass demonstrates further reduction in size compared to the previous study and now measures 2.2 cm x 2.1 cm (previously 3.5 cmx 3 cm). The previously demonstrated ipsilateral pulmonary nodule is unchanged in size in appearance. No new nodules demonstrated.

The previously demonstrate mediastinal lymphadenopathy has slightly increased in size in maximum measuring some 1.2cm in short axis in the right hilum (previously 9mm) cm).

Abdomen/Pelvis
No focal hepatic lesions. The spleen, pancreas, adrenals, kidneys are unremarkable.
No underlying bony lesions.

Comment:
The appearance are in keeping with oligoprogressive disease where the right upper lobe lesion has reduced in size and now measures 2.2x2.1 cm.
There is however the increase in the size of the right ipsilateral hilar lymph node that there is a 1.4 cm in short axis.

*this is a mock patient case prepared by a consultant radiologist for the purposes of the simulation sessions conducted by the Institute of Global Health Innovation, Imperial College London

NHS
Imperial College Healthcare
NHS Trust*

**IMAGING REPORT**

BLOGGS, JOSPEH

IGHI005

**RYJ20526266 30/06/2020**
CT Chest/Abdomen/Pelvis with contrast

Clinical details:
The 64 yrl male. Diagnosis of T3 N1 M1a lung adenoca. Nov 2019 treated with Pembrolizumab monotherapy initially q3w then q6w with good tolerance. Now has deranged ALT. ? mets

.
Report:
Contrast enhanced axial images of the chest abdomen and pelvis.
Comparison is made with a previous CT scan of Feb 2020 and 2nd June 2020.

Chest
The previously demonstrated right upper lobe lesion has unfortunately increased in size and now measures 6.5cm x 6.5 cm (previously 4 x 5 cm).
There is evidence or some subsegmental atelectasis surrounding this lesion. There are two new ipsilateral lung nodules in the right upper lobe.

The right hilar lymph node is increased in size and now measures 2.5 cm short axis.
Extensive lymphadenopathy is now demonstrated.

SAMPLE

Abdomen/Pelvis

Since the previous scan there is now evidence of 2 new contrast enhancing lesions within the right and left hepatic lobe measuring 1.2 and 1.6 cm respectively.
No evidence of abdominal or pelvic lymph node enlargement.
The adrenals are unremarkable. The spleen, pancreas, gallbladder and kidneys are unremarkable. No evidence of bony lesions.
The unprepared bowel is unremarkable.

Comment:
Appearances are in keeping with progressive disease with increase in the size of the right upper lobe lesion, increase in the size and extent of mediastinal lymphadenopathy.
Two new hepatic lesions within the right and left lobes.

The clinician has been emailed with the findings

*this is a mock patient case prepared by a consultant radiologist for the purposes of the simulation sessions conducted by the Institute of Global Health Innovation, Imperial College London

Radiology reports for one of the synthetic patient cases which was developed by a consultant interventional radiologist and reverse-engineered to match to an existing lung cancer clinical trial identified by the research team at Imperial College London.

### Screenshots of NAVIFY Clinical Trial Match application (v1.4) during the simulation sessions

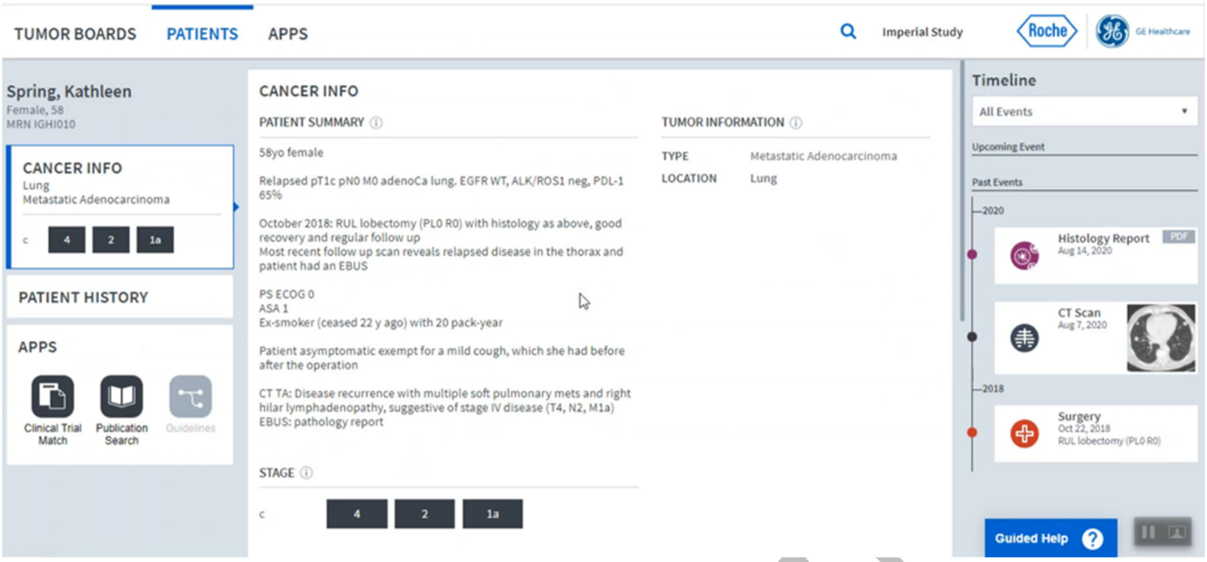


1) Overview of a patient record in NAVIFY Tumor Board, including a timeline of investigations and patient summary.

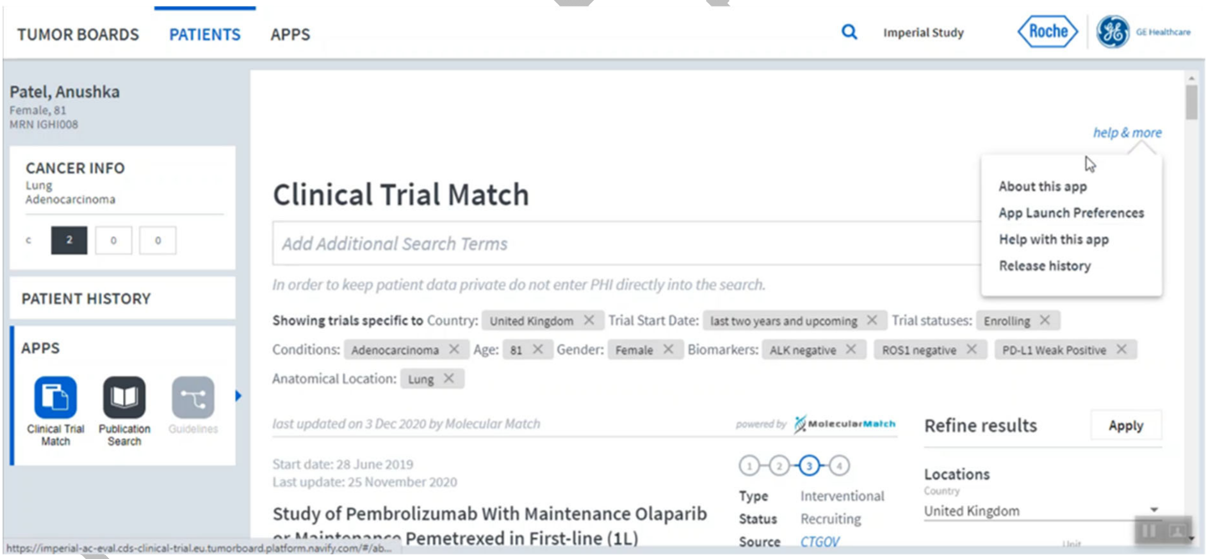


2) Overview of search results layout in NAVIFY Clinical Trial Match, including the search terms extracted from the patient record in NAVIFY Tumor Board.

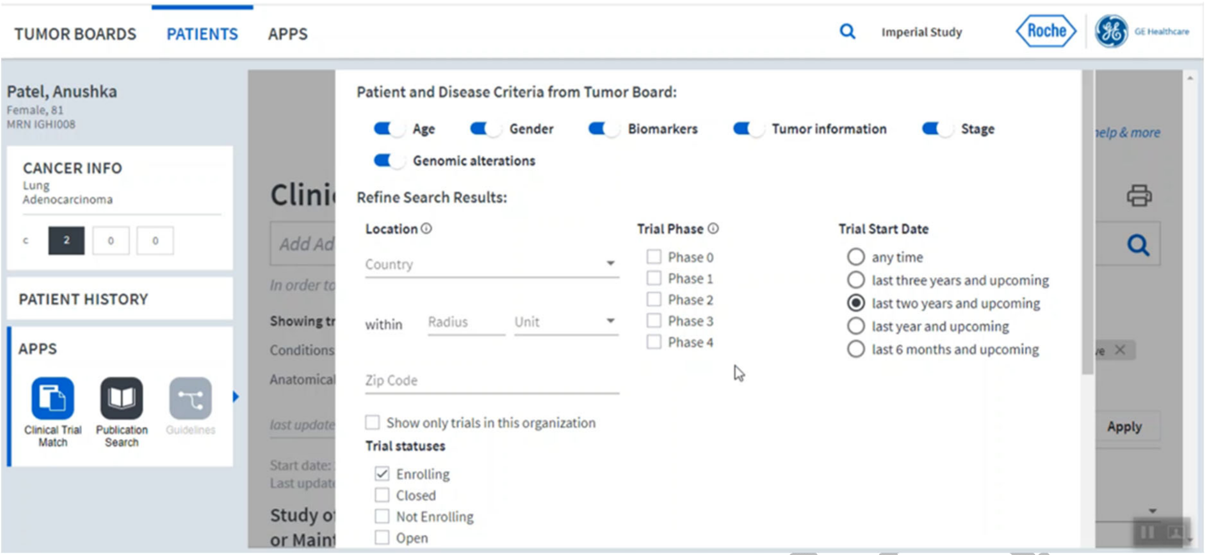


3) App Launch Preferences in NAVIFY Clinical Trial Match.

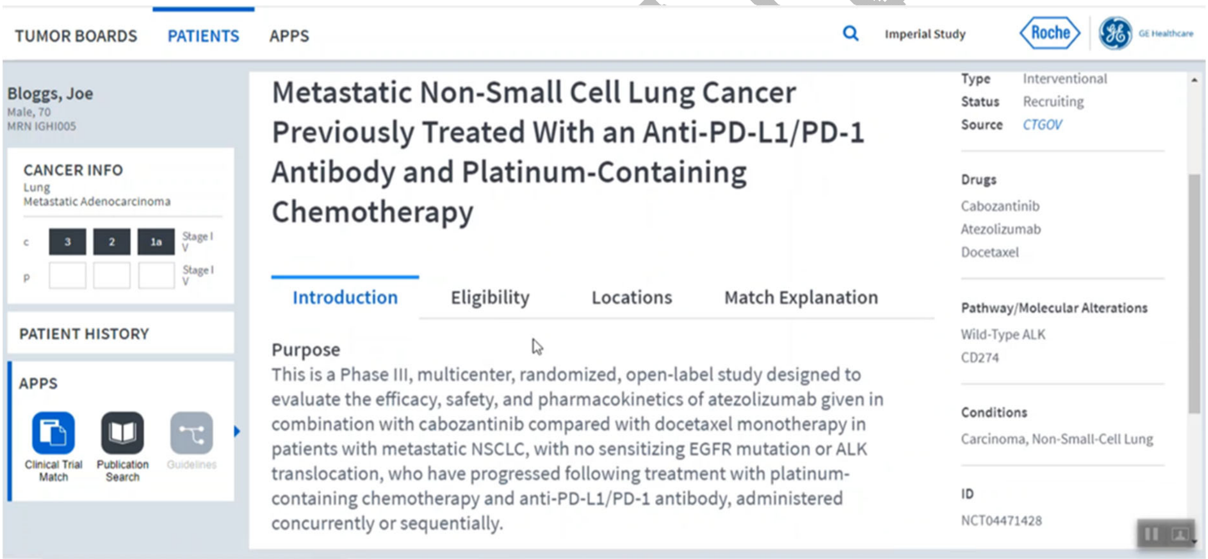


4) Information on each of the matching clinical trials is extracted from the relevant database(s) and presented in NAVIFY Clinical Trial Match.

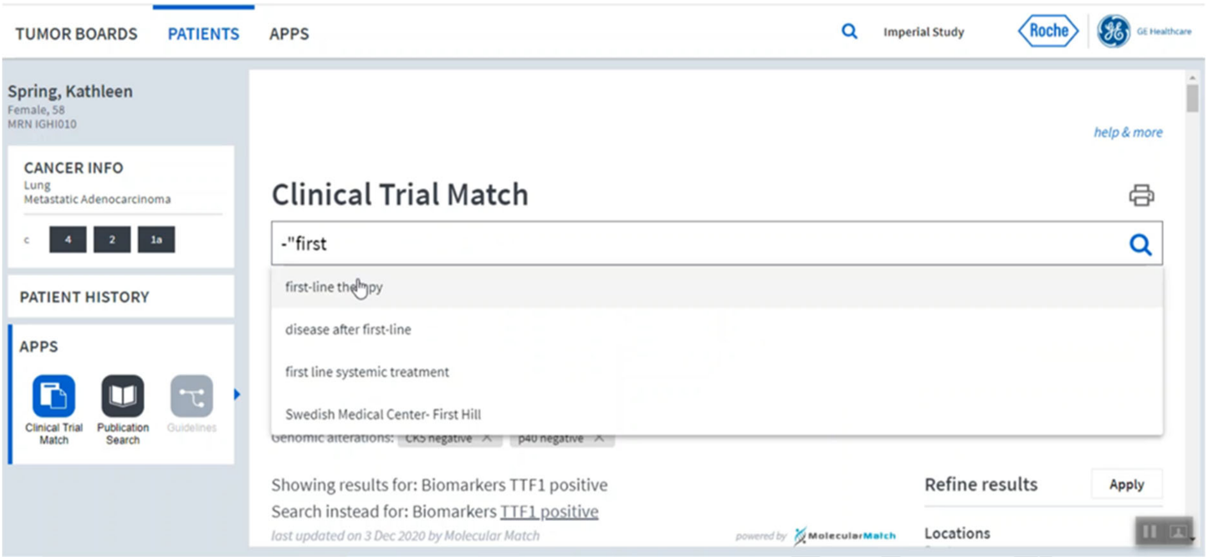


5) Example of a participant adding their own search terms in NAVIFY Clinical Trial Match.

Pre-Print Article
